# Dysregulation of the PRUNE2/PCA3 genetic axis in human prostate cancer: From experimental discovery to validation in two independent patient cohorts

**DOI:** 10.1101/2022.10.25.22281522

**Authors:** Richard C. Lauer, Marc Barry, Tracey L. Smith, Andrew Maltez Thomas, Jin Wu, Ruofei Du, Ji-Hyun Lee, Arpit Rao, Andrey S. Dobroff, Marco A. Arap, Diana N. Nunes, Israel T. Silva, Emmanuel Dias-Neto, Isan Chen, Dennis J. McCance, Webster K. Cavenee, Renata Pasqualini, Wadih Arap

**Author notes:** **Authorship notes:**. RCL and MB contributed equally to this work. Author order was determined based on order of joining the project. RP and WA jointly supervised the project. AMT: Department of Cellular, Computational and Integrative Biology, University of Trento, Trento, Italy. **Address correspondence to:** Wadih Arap, Rutgers Cancer Institute of New Jersey, Newark, 205 South Orange Avenue, New Jersey 07101, USA.

## Abstract

**BACKGROUND:** We have previously shown that the long non-coding (lnc)RNA *prostate cancer associated 3* (*PCA3*; formerly *prostate cancer antigen 3*) functions as a trans-dominant negative oncogene by targeting the previously unrecognized prostate cancer suppressor gene *PRUNE2* (a homolog of the *Drosophila prune* gene), thereby forming a functional unit within a unique allelic locus in human cells. Here we investigated the *PCA3*/*PRUNE2* regulatory axis from early (tumorigenic) to late (biochemical recurrence) genetic events during human prostate cancer progression.

**METHODS:** The reciprocal *PCA3* and *PRUNE2* gene expression relationship in paired prostate cancer and adjacent normal prostate was analyzed in two independent retrospective cohorts of clinically-annotated cases post-radical prostatectomy: a single-institution discovery cohort (n=107) and a multi-institution validation cohort (n=497). We compared the tumor gene expression of *PCA3* and *PRUNE2* to their corresponding expression in the normal prostate. We also serially examined clinical/pathological variables including time to disease recurrence.

**RESULTS:** We consistently observed increased expression of *PCA3* and decreased expression of *PRUNE2* in prostate cancer compared with the adjacent normal prostate across all tumor grades and stages. However, there was no association between the relative gene expression levels of *PCA3* or *PRUNE2* and time to disease recurrence, independent of tumor grades and stages.

**CONCLUSIONS:** We concluded that upregulation of the lncRNA *PCA3* and targeted downregulation of the protein-coding *PRUNE2* gene in prostate cancer could be early (rather than late) molecular events in the progression of human prostate tumorigenesis but are not associated with biochemical recurrence. Further studies of PCA3/PRUNE2 dysregulation are warranted.

**FUNDING:** We received support from the Human Tissue Repository and Tissue Analysis Shared Resource from the Department of Pathology of the University of New Mexico School of Medicine and a pilot award from the University of New Mexico Comprehensive Cancer Center. RP and WA were supported by awards from the Levy-Longenbaugh Donor-Advised Fund and the Prostate Cancer Foundation. EDN reports research fellowship support from the Brazilian National Council for Scientific and Technological Development (CNPq), Brazil, and the Associação Beneficente Alzira Denise Hertzog Silva (ABADHS), Brazil. This work has been funded in part by the NCI Cancer Center Support Grants (CCSG; P30) to the University of New Mexico Comprehensive Cancer Center (CA118100) and the Rutgers Cancer Institute of New Jersey (CA072720).

## Introduction

Prostate cancer is the most common cancer and the second most common cause of cancer death in men^1^, and there continues to be a pressing need for new diagnostic and therapeutic approaches for this disease, as well as better prognostic biomarkers to guide treatment. Long non-coding RNA (lncRNA) species are increasingly recognized as having regulatory functions in tumorigenesis, and nucleic acid-based therapeutics are being developed as a promising means of targeting pathogenic lncRNAs^2^. Several lncRNAs have recently been found to associate with prostate cancer, and the best known of these, *prostate cancer associated 3* (*PCA3*; formerly *prostate cancer antigen 3*) has been used clinically for many years as the most specific diagnostic biomarker for prostate cancer^3,4^; however, its prognostic significance remains uncertain. Strikingly, *PCA3* emerged first only in mammals, with further evolution in primates^5^, and, given aspects of the sequence and genomic organization, we have hypothesized that it might have been introduced into the genome by an ancient oncogenic virus^6^. In humans, *PCA3* has an unusual genomic organization, being present in an antisense direction within an intron of the protein-coding gene *PRUNE2*. Somewhat surprisingly for a molecule that is well-established as a Food & Drug Administration (FDA)- and European Medical Agency (EMA)-approved biomarker, relatively little was known about the biological function of *PCA3* until recently. Ferreira et al. showed that *PCA3* is androgen-regulated and that it promotes prostate cancer cell survival^7^. Subsequently, we have established that *PCA3* downregulates the expression of *PRUNE2* in a rather unusual way: at the RNA level by RNA editing mediated via adenosine deaminase RNA specific (ADAR)-family members^8^. We have showed that expressing ectopic *PCA3* or, alternatively, silencing *PRUNE2* induced cell transformation and cell proliferation *in vitro*, increased adhesion and migration of prostate cancer cells, and yielded larger tumors in xenograft tumor models. The opposite biological effects were seen with *PCA3* silencing or ectopic *PRUNE2* expression^8^. Preliminary studies of human prostate cancer samples compared to normal prostate showed increased *PCA3* expression, decreased *PRUNE2* expression, and evidence for RNA editing of these genes. Based on these experimental findings, we proposed that there is a functional molecular axis in human prostate cancer in which *PCA3* acts as a transdominant-negative oncogene to downregulate a previously unrecognized tumor suppressor gene, *PRUNE2*^8^.

Here we propose that this molecular interplay may serve as a translational target for diagnostic and/or therapeutic intervention in human prostate cancer. First, we present additional correlative evidence from two retrospective post-surgical primary prostate cancer cohorts in support of our experimental model of *PCA3* as a dominant-negative oncogene and *PRUNE2* as a tumor suppressor gene, and for their co-regulation in human prostate cancer. Moreover, we examine the dysregulation of the *PCA3*/*PRUNE2* regulatory axis across tumors of different grades (patterns), stages, and groups^9,10^. Finally, we assess whether tumor expression levels of *PCA3* and/or *PRUNE2* are prognostic of biochemical disease recurrence after surgery.

## Methods

### Discovery patient cohort

Based on a power analysis using gene expression data from our prior work^8^, for the UNMCCC single-institution discovery cohort, we searched the archives of the Department of Pathology at the UNM School of Medicine for at least 100 consecutive patients (final cohort size: n=107) who had a radical prostatectomy as the primary treatment for organ-confined prostate cancer between the years 2001 and 2013 and who had the following clinical and pathological attributes: final post-prostatectomy Gleason Score 7 [either Gleason Group 2 (3+4) or Gleason Group 3 (4+3)], pathologic stage pT2 or pT3a, negative surgical margins, negative for seminal vesicle invasion, no evidence of local or distant metastasis, and no prior treatment for prostate cancer. The following additional data were retrospectively abstracted from the individual medical records: age at surgery, race, presence of recurrence, type of recurrence (i.e., biochemical, local, metastatic), and disease-free survival time. Biochemical disease recurrence was defined as a detectable serum prostate-specific antigen (PSA) concentration of at least 0.2 ng/mL post-operatively. Lost to follow up was defined as not having been followed up at the UNMCCC after their urological surgery. All included cases had an independent pathologic re-review by a Board-certified pathologist with expertise in urological oncology (MB), with confirmation of diagnosis, Gleason-based analysis (grading, scoring, and grouping), standard TNM staging, and margin status post-resection. A small number of identified cases (<5%) had to be excluded due to the very limited amount of tumor present.

### Microdissection of tumor and normal prostate (nonneoplastic prostatic glandular tissue) for the discovery cohort

To obtain tumor for RNA analysis, a representative carcinoma-containing formalin-fixed paraffin embedded (FFPE) block was chosen from each case. Contiguous foci of tumor were marked on the glass slide such that the density of tumor cells was at least 75%. The boundary of the corresponding areas on the tumor block was scored with a blade tip, effectively allowing microdissection of tumor in the process of microtome sectioning. Multiple 10μm sections were cut, depending on the area of the tumor focus/foci. In 24 (22.4%) of the cases, we also microdissected areas of nonneoplastic prostatic glandular tissue away from tumor in a similar manner, again also aiming for at least 75% epithelial density.

### Measurement of PRUNE2 and PCA3 gene expression in the discovery cohort by quantitative RT-PCR

Briefly, gene expression for *PCA3* and *PRUNE2* were determined by quantitative reverse transcription polymerase chain reaction (qRT-PCR) by using TaqMan gene expression assays (Thermo Fisher Scientific) with amplicon detection via a LightCycler 96 (Roche Diagnostics). Gene expression was quantified by the relative logarithmic RT-PCR threshold cycles (ΔCt) between the target genes and housekeeping control genes^11^. Specifically, total RNA was extracted from the microdissected FFPE sections using the PureLink™ FFPE Total RNA Isolation Kit (Thermo Fisher Scientific, Cat. No. K1560-02). RNA was quantified on a NanoDrop ND-1000 Spectrophotometer (Thermo Fisher Scientific), and the average A260/A280 ratio was 1.94 (range 1.88 to 2.07), indicating optimal quality of the RNA extracted for gene expression assays. RNA was then further quantified with the Qubit® RNA HS Assay Kit (Thermo Fisher Scientific, Cat. No. Q32852) on a Qubit 2.0 (Thermo Fisher Scientific) for accurate RNA concentration. RNA integrity was evaluated with the Agilent RNA 6000 Nano kit (Agilent Technologies, Cat. No. 5067-1511) on an Agilent 2100 Bio analyzer (Agilent Technologies). To remove genomic DNA contamination, RNA samples were treated with 2U of DNase I (Thermo Fisher Scientific, Cat. No. 18068-015) per 2μg of total RNA. All procedures were performed according to the manufacturer’s standard protocols.

Reverse Transcription was performed in triplicate in order to create enough cDNA for the entire project. 500ng RNA in each of three tubes was reverse transcribed with the High-Capacity RNA-to-cDNA™ Kit (Thermo Fisher Scientific, Cat. No. 4387406) in a final volume of 20μl, according to the manufacturer’s instructions. Reverse transcription was carried out in a Gene Amp PCR System 9700 (Applied Biosystems) at 37°C for 60 minutes and terminated by 95°C for 5 minutes. Then three aliquots were combined for the following experiments.

For the Thermo Fisher Scientific TaqMan gene expression assay experiments, three (Hs00322421_m1, Hs00999960_m1, and Hs01060890_m1) and two (Hs01371939_g1 and Hs03462121_m1) assays were chosen for target genes *PRUNE2* and *PCA3* respectively, (designated PR1, PR2 and PR3, and PC1 and PC2). Three endogenous controls GAPDH (Hs02758991_g1), HPRT1 (Hs02800695_m1), and UBC (Hs01871556_s1) were selected (designated C1, C2 and C3)^12^. Each *PRUNE2* assay and *PCA3* assay was labeled with FAM and paired with a VIC labeled endogenous control in a duplex reaction, with separate reactions to include all of the three endogenous controls. Therefore, a total of fifteen duplex gene expression mixes, nine for *PRUNE2* and six for *PCA3*, was required for all specimens (**Fig. 1 - Source Data File 1** [tumor] and **Fig. 1 - Source Data File 2** [normal]).

**Figure 1:**
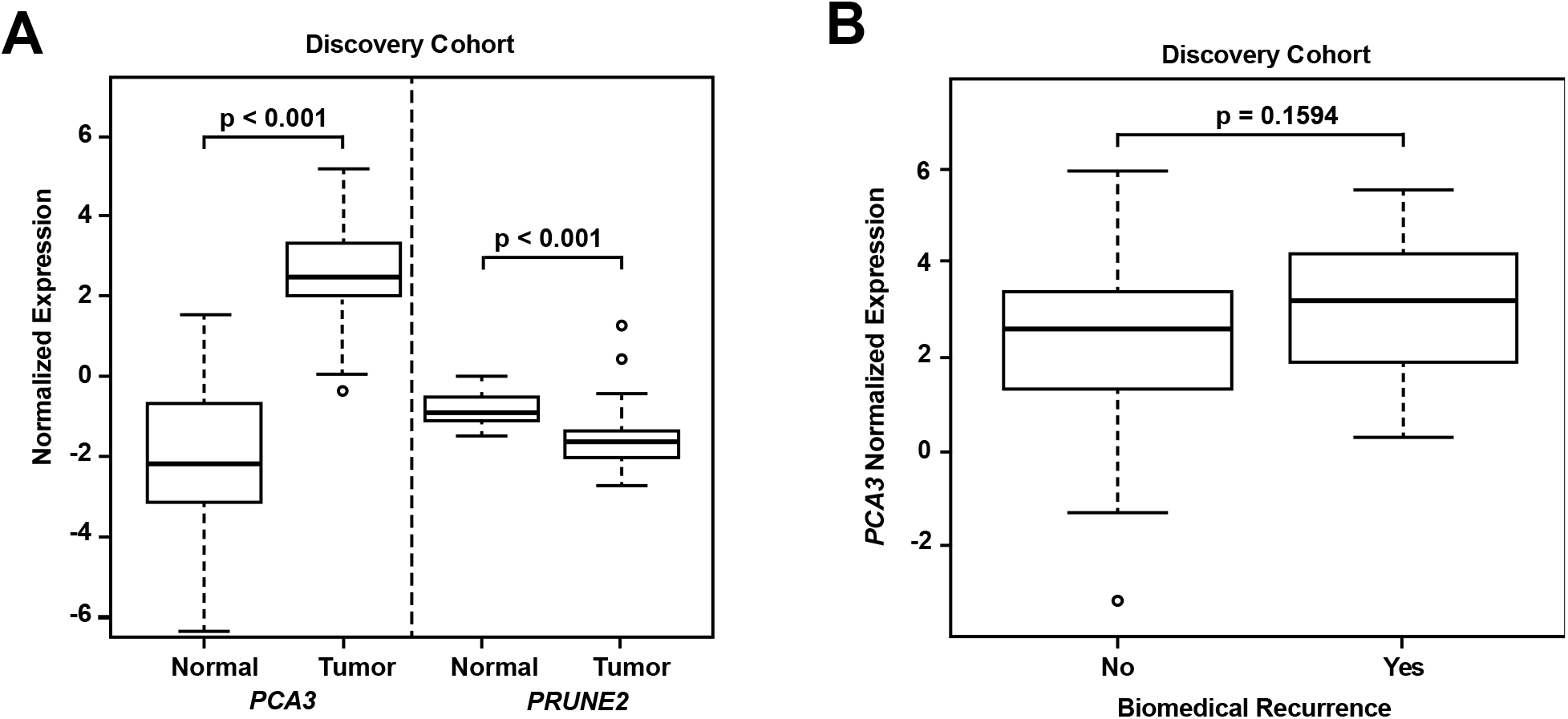
Analyses of discovery prostate cancer cohort. **(A)** *PRUNE2* and *PCA3* expression in tumor (**Fig. 1 - Source Data File 1**) and nonneoplastic (**Fig. 1 - Source Data File 2**) prostatic samples. Calculated values available in **Fig. 1 - Source Data File 3. (B)** Tumor *PCA3* expression by biochemical recurrence status. *PCA3* expression in patients without versus with biochemical recurrence in the discovery cohort. No significant difference in median expression was seen in this cohort. Box plots of gene expression (normalized expression) in the discovery cohort. The horizontal line within each box represents the median value, while the box represents the interquartile range, and the whiskers extend out to 1.5 times the interquartile range. Outliers are represented by circles. P-values are noted for the indicated comparisons.

Each duplex gene expression assay was then performed in triplicate for all specimens following manufacturer’s standard protocols, for a total of 45 expression measures for each case. qRT-PCR was performed with the TaqMan Gene Expression Master Mix (Thermo Fisher Scientific, Cat. No. 4369514) using 1μl of each TaqMan target gene assay (20×FAM) and endogenous controls assay (20×VIC), 1μl of cDNA template (equivalent to 25ng RNA input), and 7μl of RNase-free water for a 20μl final reaction mixture. A non-template control was included in every master mix in every 96-format tray. In addition, in order to evaluate inter-plate variation, we also included one RNA sample, in triplicate, in all the 96 format trays. Analysis of these controls indicated that there were no significant batch effects (data not shown). The qRT-PCR product detection was achieved on a LightCycler 96 (Roche Diagnostics). The cycle program was: at 95°C for 10 minutes, followed by 40 cycles at 95°C for 15 seconds and at 60°C for 1 minute. Quantification of target and control genes (Cq) in each sample was performed by LightCycler® 96 SW 1.1 (Roche Diagnostics).

### Validation patient cohort

For the TGCA patient validation cohort (n=497 patients), we first downloaded clinical data along with the expression of the lncRNA *PCA3* and the *PRUNE2* gene (http://cancergenome.nih.gov) with the UCSC Xena browser^13,14^, together with paired nonneoplastic samples in 52 of the cases (10.5%). The following clinical and pathological characteristics were included in the study: age at diagnosis, vital status, tumor Gleason-based analysis (grading, scoring, grouping), pathological stage, status of biochemical recurrence, and time to recurrence. Gene expression was calculated with RSEM^15,16^. By using the available dataset, we evaluated *PCA3* and *PRUNE2* gene expression values in terms of tumor versus nonneoplastic prostate, biochemical recurrence, pathologic T stage, Gleason analysis (Grade, Score, and Group), age at pathology-proven diagnosis. Because the regulation of *PRUNE2* by *PCA3* occurs at the RNA level by the formation of an RNA hetero-duplex, we also evaluated the ratio of the expression of the two genes in terms of the clinical and pathological variables for each patient of the cohort.

### Statistics

Demographic and clinical variables were summarized with descriptive statistics. For the discovery cohort, the mean and median of gene expressions across multiple control genes and assays were summarized, and these were used as measures for gene expression of *PRUNE2* and *PCA3* relative to endogenous housekeeping controls for each case. More detailed methods are described in the supplemental materials.

Testing for differences of *PCA3* and *PRUNE2* expression between paired tumor and nonneoplastic prostate expression was by the Wilcoxon signed rank test. The Kruskal-Wallis test was used when comparing three or more groups. Assessment for significant differences of gene expression by recurrence status was by Wilcoxon rank sum test. The Kaplan-Meier product limit method with log-rank test was used to explore the relationship between gene expression levels or the ratio and the time to recurrence. Multivariable Cox proportional hazard modeling was used to fit for the association between time to recurrence and expression levels of *PRUNE2* or *PCA3* or their ratio, while controlling for multiple clinical covariates. All statistical analyses were carried out by using the SAS (9.4) or R software package (R 3.4.5), unless otherwise indicated (R and SAS codes are available in the **Source Code File**).

### Study approval

For the discovery cohort, there was University of New Mexico Health Sciences Institutional Review Board (IRB) approval (HRRC15-138), and the study was carried out in accordance with the United States Common Rule.

## Results

### Discovery Single-Institutional Cohort

In the initial single-institution discovery cohort from the University of New Mexico Comprehensive Cancer Center (UNMCCC), patients with intermediate-risk (Gleason Score 7; corresponding to Gleason Groups 2 and 3) organ-confined prostate cancer (n=107) met the criteria for inclusion in this study (**Table 1**). Briefly, the mean age of the cohort was 63 years old (ranging from 45 to 84 years old); most patients (85%) were non-Hispanic white, but Hispanic (7.5%), American Indian/Native American (2.8%), and African American (2.8%) men were also represented. All patients had final Gleason Score 7 adenocarcinoma after radical prostatectomy, with 86.9% being 3+4=7 (Gleason Group 2) and 13.1% being 4+3=7 (Gleason Group 3). The pathologic stage distribution was as follows: 74.8% were pT2 and 25.2% were pT3a. Nineteen of the patients (17.8%) had biochemical recurrence discovered during follow up, including one with documented local recurrence and one with documented metastases. Five patients (4.7%) were lost to follow up.

**Table 1.**
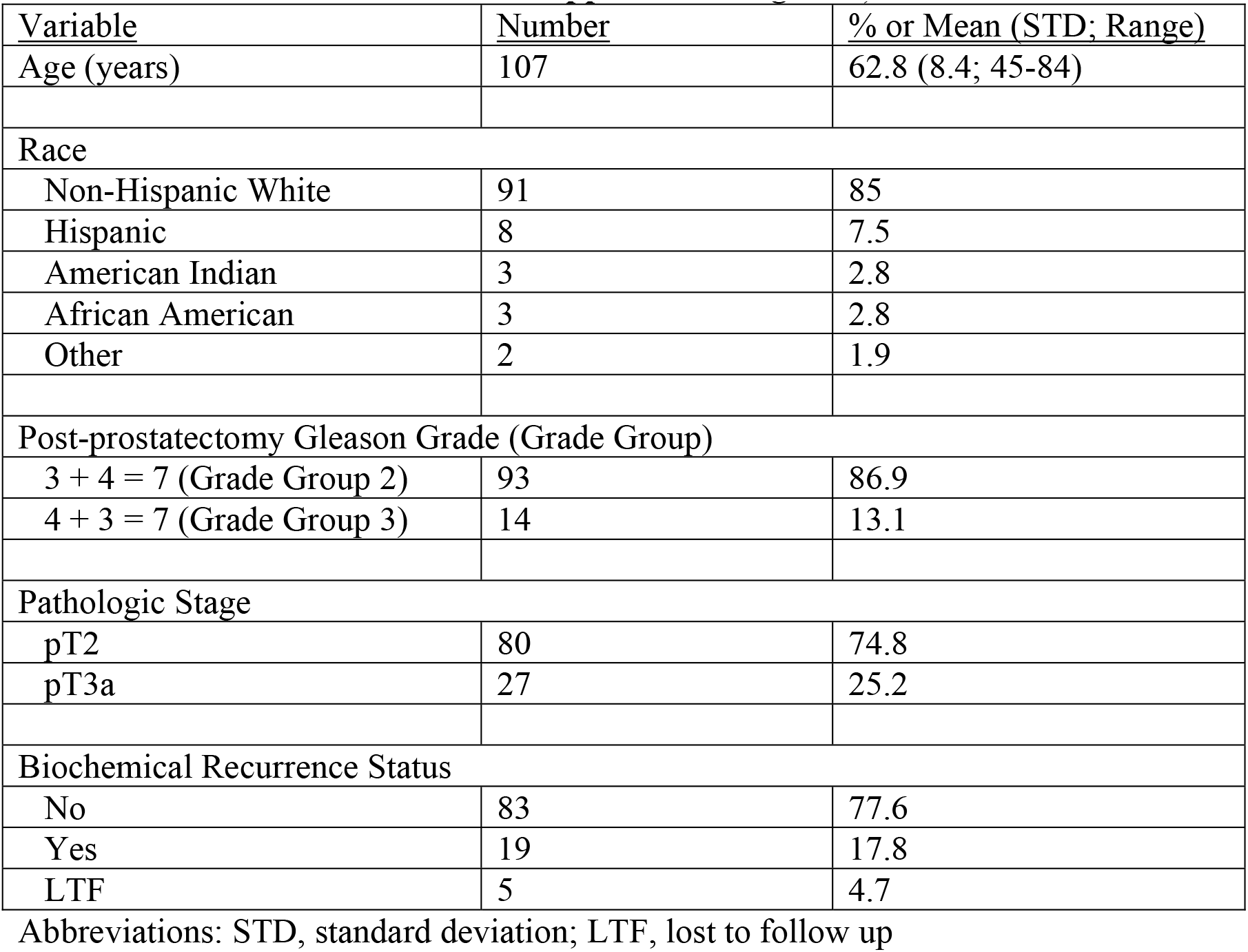
Discovery Cohort: clinicopathologic features of the 107 patients in this study who had radical prostatectomy at UNM for localized prostatic adenocarcinoma (Further details in **Table 1 – Source Data File 1** and **Table 1 – Supplemental Figure 1**)

| Variable | Number | % or Mean (STD; Range) |
| --- | --- | --- |
| Age (years) | 107 | 62.8 (8.4; 45-84) |
| Race |  |  |
| Non-Hispanic White | 91 | 85 |
| Hispanic | 8 | 7.5 |
| American Indian | 3 | 2.8 |
| African American | 3 | 2.8 |
| Other | 2 | 1.9 |
| Post-prostatectomy Gleason Grade (Grade Group) |  |  |
| 3 + 4 = 7 (Grade Group 2) | 93 | 86.9 |
| 4 + 3 = 7 (Grade Group 3) | 14 | 13.1 |
| Pathologic Stage |  |  |
| pT2 | 80 | 74.8 |
| pT3a | 27 | 25.2 |
| Biochemical Recurrence Status |  |  |
| No | 83 | 77.6 |
| Yes | 19 | 17.8 |
| LTF | 5 | 4.7 |

Abbreviations: STD, standard deviation; LTF, lost to follow up

RNA extraction and quantitative real-time polymerase chain reaction (qRT-PCR) were successful in all microdissected tumor samples (n=107). In 24 of these cases (22.4%), we extracted RNA from benign prostatic glandular tissue away from tumor (hereafter termed “normal prostate”: qRT-PCR was successful in all cases for *PRUNE2* (n=24, 100%) and in most cases for *PCA3* (n=21, 87.5%). Comparing *PRUNE2* and *PCA3* expression in prostatic adenocarcinoma with expression in normal prostate (all relative to endogenous housekeeping controls), we found consistent trends for both genes in multiple assays, with lower expression of *PRUNE2* in tumor as compared with normal prostate and higher expression of *PCA3* in tumor as compared with normal prostate (**Fig. 1 - Source Data File 3**). These results are summarized in **Figure 1A** and as follows. Relative to controls, *PCA3* expression was significantly higher in prostatic adenocarcinoma (mean: 2.46; standard deviation: 1.28) than normal prostate (-1.99; 2.63, [p-value<0.001]). Relative to controls, *PRUNE2* expression was significantly lower in tumor (mean: -1.48; standard deviation: 0.92) than normal prostate (-0.78; 0.4, [p-value<0.001]).

We next explored the association between biochemical recurrence and tumor expression levels of *PRUNE2, PCA3* and the ratio of *PRUNE2* to *PCA3* expression by using several approaches. First, we compared the gene expression values and their ratio by recurrence status. In patients who recurred compared to those who did not, we found no significant difference in mean expression values of *PRUNE2* (-1.6 vs. -1.58; p-value=0.68), *PCA3* (2.98 vs. 2.43; p-value=0.16), or their ratio (-1.61 vs. -1.21, p-value=0.48). The different expression levels by recurrence were not significant (**Figure 1B, Supplemental Figure S1**). Next, for *PRUNE2* expression, *PCA3* expression, and their ratio, we regrouped the cancer cases according to whether the values were greater than (deemed “high”) or less than/equal to (deemed “low”) their respective mean values. By using the Kaplan-Meier product limit methodology and the log-rank test, we found no significant associations between high or low levels and time to recurrence for *PRUNE2* expression (p-value=0.24), *PCA3* expression (p-value=0.22), (**Figure 2** and **Tables 2-3**), or their ratio (p-value=0.84). As a further assessment of association between gene expression and time to biochemical recurrence, we used Cox proportional hazards modeling and found no significant associations of time to biochemical recurrence with expression of *PRUNE2* (hazard ratio [HR]: 0.97; 95% confidence interval [CI]: 0.58–1.63, [p-value=0.91]) or *PCA3* (HR: 1.21; 95% CI: 0.91–1.6, [p-value=0.19]), or their ratio (HR: 0.99; 95% CI: 0.92–1.1, [p-value=0.82]). By multivariable Cox modeling, we did not find that expression of *PRUNE2, PCA3* or their ratio added any additional predictive information for recurrence to that provided by clinical or pathological variables, as presented in supplemental results and **Supplemental Table S1**.

**Table 2.** Number of patients at risk over time (see **Figure 2A**)

|  | Years Post Radical Prostatectomy |  |  |  |
| --- | --- | --- | --- | --- |
| *PRUNE2* Expression | 0 | 5 | 10 | 15 |
| High | 51 | 15 | 5 | 0 |
| Low | 56 | 14 | 2 | 0 |

**Table 3.** Number of patients at risk over time (see **Figure 2B**)

|  | Years Post Radical Prostatectomy |  |  |  |
| --- | --- | --- | --- | --- |
| *PCA3* Expression | 0 | 5 | 10 | 15 |
| High | 59 | 16 | 2 | 0 |
| Low | 48 | 13 | 5 | 0 |

**Figure 2.**
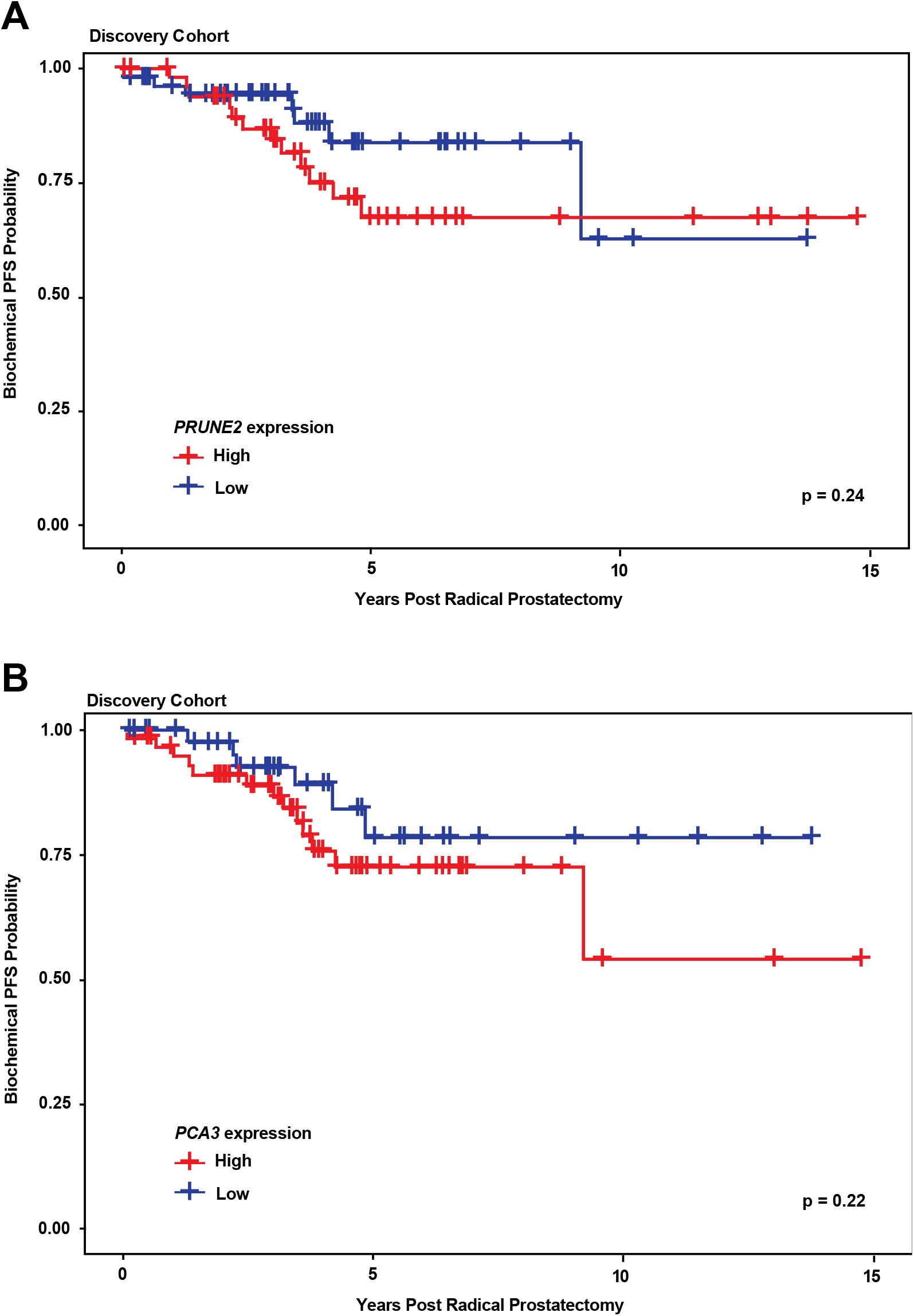
Kaplan-Meier curves illustrating time to event (biochemical recurrence) among patients post-prostatectomy from the discovery cohort, stratified by “high” gene expression (greater than mean expression, red line) versus “low” gene expression (less than or equal to mean expression, blue line), for (**A**) tumor *PRUNE2* expression, and (**B**) tumor *PCA3* expression. There is was no significant association of high versus low expression levels and time to recurrence by log rank testing for either *PRUNE2* or *PCA3*.

### Validation/Confirmation Multi-Institutional Cohort

For the analysis of a second multi-institutional validation/confirmation and expansion prostate cancer cohort from The Cancer Genome Atlas (TCGA), patient clinical data (**Table 4**) and gene expression data were available on men with prostate cancer (n=497). Gene expression data from nonneoplastic prostatic tissue (hereafter termed “normal prostate”) were also available in 52 (10.5%) of the cases. The basis for the cohort has been described previously^13^. Briefly, the cohort comprised men whose ages ranged from 41 to 78 years old, and who had a radical prostatectomy for primary prostate cancer. The distribution of prostate cancer grades was as follows: Gleason Score 3+3=6 (Group 1), 9.0%; Gleason Scores 3+4=7 or 4+3=7 (Groups 2 and 3), 49.7%; Gleason Score 4+4=8 (Group 4), 12.9%; and Gleason Scores 4+5=9 or 5+4=9, 27.6%; and 5+5=10 (Group 5), 0.8%. For pathologic stage, most tumors were pT2c (33%), pT3a (32%), or pT3b (27%), with a small fraction being pT4 (2%). Data on disease recurrence were available for 429 men (86.3%), with 58 (11.7% [13.5% of those with follow-up information available]) having biochemical recurrence.

**Table 4.** Validation cohort: clinicopathologic features of the 497 patients in the prostate cancer TCGA data set, with a total of 549 tissue samples included.

| Variable | Number | % or Mean (STD; Range) |
| --- | --- | --- |
| Age (years) | 497 | 61 (6.8; 41-78) |
| Vital Status |  |  |
| Alive | 487 | 97.9 |
| Dead | 10 | 2.1 |
| Sample Type |  |  |
| Primary tumor | 497 |  |
| Normal (non-malignant) prostate | 52 |  |
| Post-prostatectomy Gleason Grade (Grade Group) |  |  |
| 6 (Grade Group 1) | 45 | 9 |
| 7 (Grade Groups 2 & 3) | 247 | 49.7 |
| 8 (Grade Group 4) | 64 | 12.9 |
| 9 (Grade Group 5) | 137 | 27.6 |
| 10 (Grade Group 5) | 4 | 0.8 |
| Pathologic Stage |  |  |
| pT2a | 13 | 2.6 |
| pT2b | 10 | 2 |
| pT2c | 164 | 33 |
| pT3a | 158 | 32 |
| pT3b | 135 | 27 |
| pT4 | 10 | 2 |
| Unknown | 7 | 1.4 |
| Biochemical Recurrence Status |  |  |
| No | 371 | 74.6 |
| Yes | 58 | 11.7 |
| Unknown | 68 | 13.7 |

Abbreviation: STD, standard deviation

We compared gene expression levels (log_2_ RNA-Seq by Expectation-Maximization [RSEM]) in prostatic adenocarcinoma to those in normal prostate samples in the data set from TCGA (**Figure 3A**): *PCA3* had significantly increased expression in carcinoma [median: 12.4; interquartile range (IQR): 10.3-13.7] as compared with normal prostate [median: 6.9; IQR: 5.2-9.6, [p-value<0.001]), and *PRUNE2* showed simultaneous lower expression in carcinoma (median: 11.4; IQR: 10.7-12.0) versus normal prostate (median: 12.2; IQR: 11.8-12.6, [p-value<0.001]). As depicted in **Figure 3B**, comparing tumor gene expression across different prostate cancer pathologic grades, *PCA3* expression was significantly lower in tumors with Gleason Score greater than 7 (median: 11.6; IQR: 8.4-13.4) than in tumors with Gleason Score 7 (median: 12.8; IQR: 11.3-13.8, [p-value<.001]) or less than 7 (median: 12.5; IQR: 11.8-13.7, [p-value=0.01]). *PRUNE2* showed a small decrease in expression in tumors with Gleason Score greater than 7 (median: 11.3; IQR: 10.4-11.9) as compared with tumors with Gleason Score 7 (median: 11.5; IQR: 10.8-12.1, [p-value=0.014]) or less than 7 (median: 11.6; IQR: 11.0-12.1, [p-value=.049]). As shown in **Figure 3C**, comparing tumor gene expression across different tumor pathologic stages, *PCA3* expression was higher in pT2 tumors (median: 12.6; IQR: 11.2-13.8) than in tumors that were pT3 (median: 12.2; IQR: 9.7-13.6, [p-value=0.01]) or pT4 (median: 12.1; IQR: 9.6-12.7, [p-value=0.61]). There was no significant difference (p-value>0.05) between *PRUNE2* expression levels between the different tumor stages: pT2 (median: 11.4; IQR: 10.7-12.1), pT3 (median: 11.3; IQR: 10.6-12.0), and pT4 (median: 11.7; IQR: 10.8-12.1). We also found that the ratio of *PCA3/PRUNE2* expression showed similar associations with Gleason Score and pathologic stage as were seen with *PCA3* expression (data not shown). Overall, despite the differences in gene expression among tumor grades and stages, the median expression of *PCA3* was significantly higher in all tumor grades and stages than the expression of *PCA3* in normal prostate, and, inversely, the median expression of *PRUNE2* in all tumor grades and stages was significantly less than the expression of *PRUNE2* in normal prostate.

**Figure 3.**
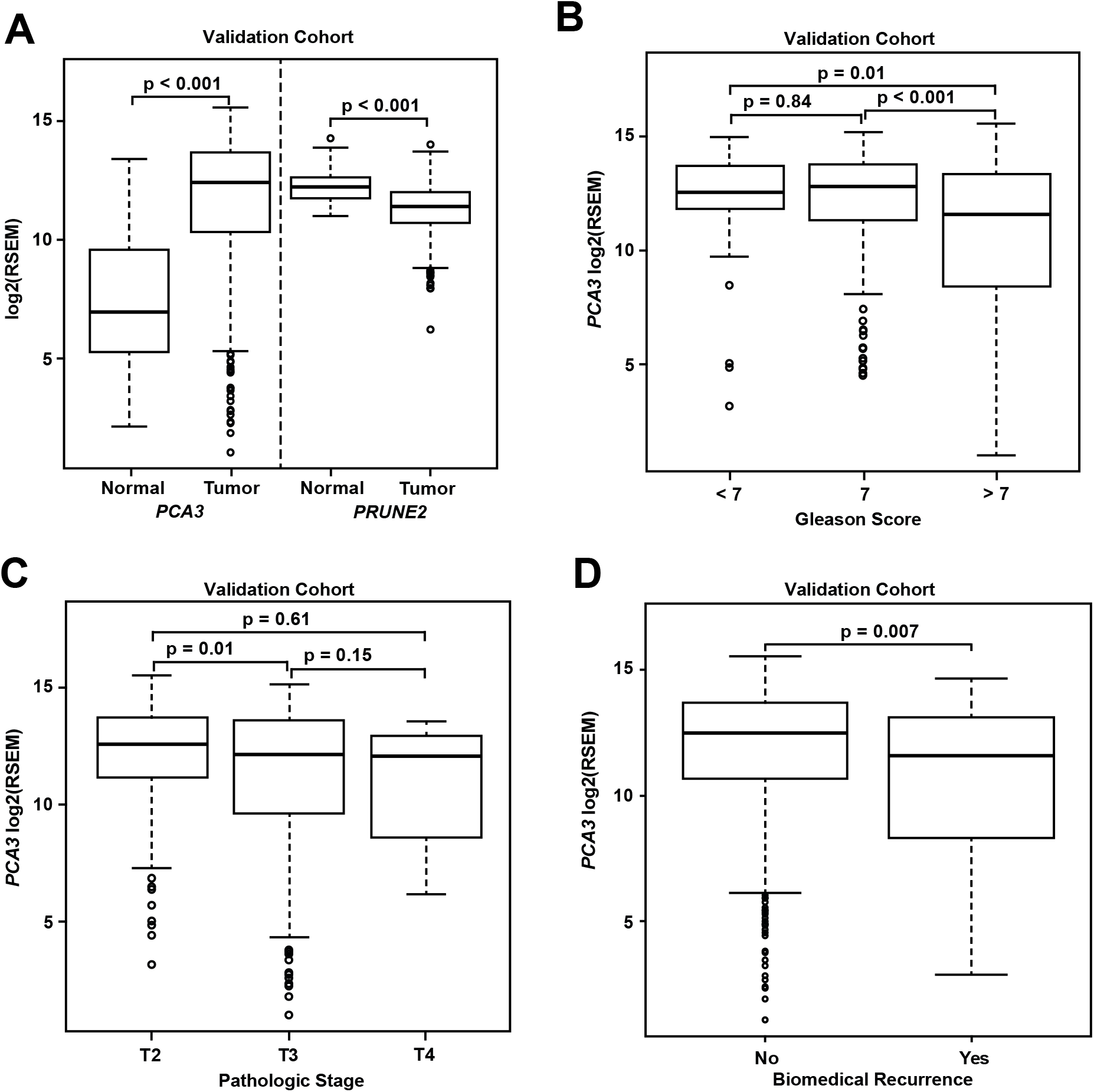
Analyses of TCGA prostate cancer validation cohort. (**A)** *PCA3* and *PRUNE2* expression in nonneoplastic prostatic glandular tissue and in prostatic adenocarcinoma in the TCGA cohort illustrating consistent gene expression differences between tumor and nonneoplastic prostate in both cohorts. (**B, C**) *PCA3* expression in the cohort from TCGA across Gleason grades (**B**) showing lower expression in higher grade (>7) tumors and across tumor stages (**C**) showing lower expression in higher stage tumors. All illustrated tumor grades and stages show higher expression than nonneoplastic prostatic epithelium. (**D**) *PCA3* expression in patients without versus with biochemical recurrence in the TCGA cohort. In the TCGA cohort, lower *PCA3* median expression was associated with biochemical recurrence. Box plots of gene expression in the TCGA cohort is reported as log2RSEM data. The horizontal line within each box represents the median value, while the box represents the interquartile range, and the whiskers extend out to 1.5 times the interquartile range. Outliers are represented by circles. P-values are noted for the indicated comparisons. (RSEM: RNA-Seq by Expectation-Maximization).

As shown for the discovery cohort, we also evaluated the relationship between PCA3 and PRUNE2 expression levels and recurrence status. We found that patients who had biochemical recurrence after prostatectomy had significantly lower tumor expression levels of *PCA3* (median, 11.58; IQR, 8.28-13.14) than those who did not recur [12.51; 10.64-13.71, (p-value<0.01); **Figure 3D**]. However, we did not see an association between tumor *PCA3* expression and biochemical recurrence on multivariable Cox proportional hazards modeling when adjusting for tumor grade, stage, and age at diagnosis [HR, 0.96; 95% CI, 0.87-1.04, (p-value=0.36)], as presented in supplemental results and **Supplemental Table S2**. We did not see a significant association between *PRUNE2* expression in those patients that had biochemical recurrence as compared with those patients who did not recur (**Supplemental Figure S2**).

## Discussion

Here we assessed the tumor and control adjacent normal prostatic glandular tissue expression of the lncRNA *PCA3* and the protein-coding *PRUNE2* gene in two independent retrospective cohorts of patients with primary organ-confined prostate cancer after treatment by radical prostatectomies (**Figure 4**). As compared with normal prostate, we found that prostate cancer showed consistent increased expression of *PCA3* and consistent decreased expression of *PRUNE2* in tumors across a broad range of pathological attributes (i.e., Gleason grades, scores, groups and stages) in both patient cohorts. These findings support the mechanistic role of a tumor-specific molecular axis in which *PCA3* acts as dominant-negative oncogene and *PRUNE2* as a tumor suppressor gene in human prostate cancer and indicate that the interplay between these genes is dysregulated early in prostate cancer.

**Figure 4.**
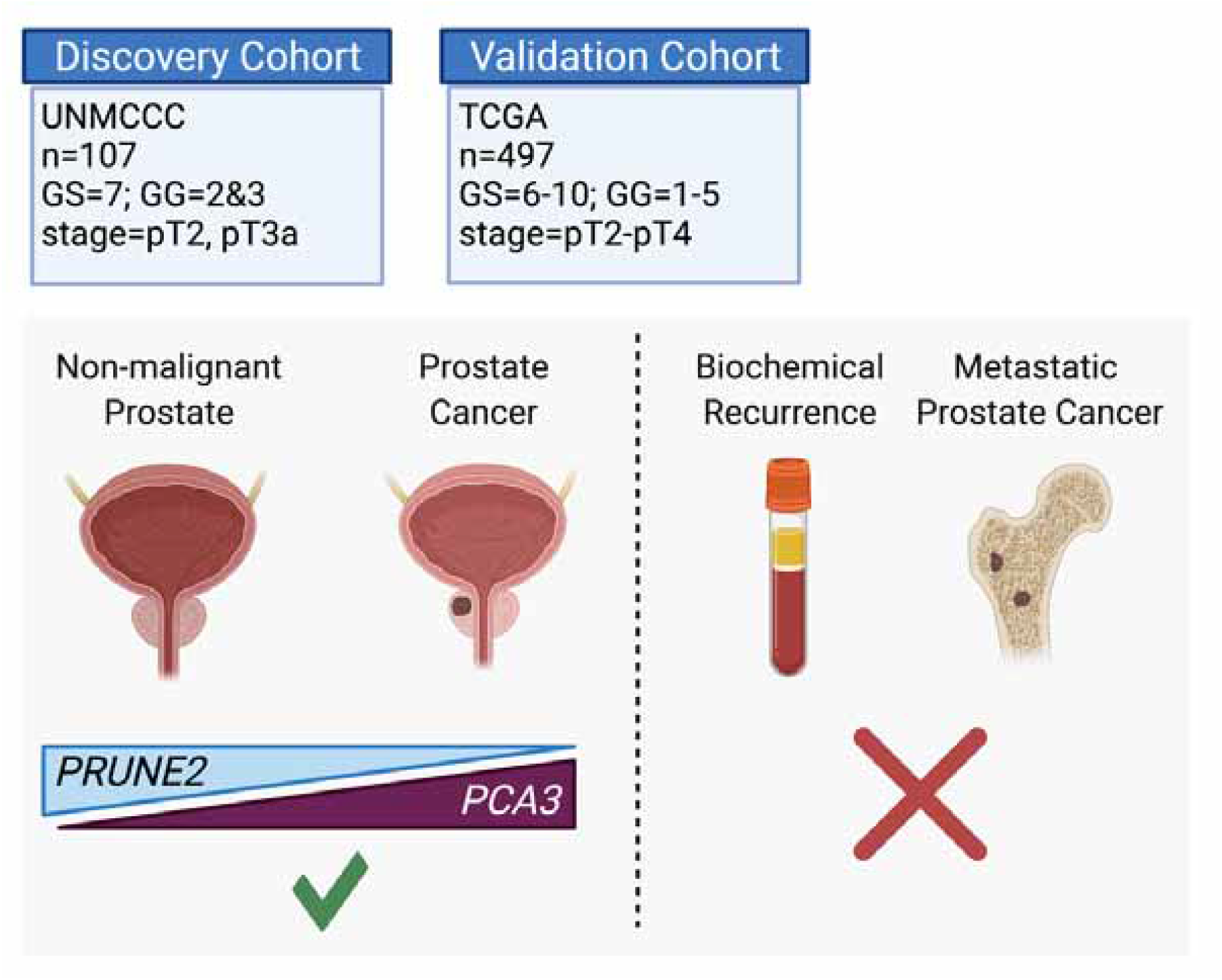
Graphical summary of the analyses. *PCA3* and *PRUNE2* gene expression relationship in paired prostate cancer and adjacent normal prostate was analyzed in two independent retrospective cohorts of clinically-annotated cases post-radical prostatectomy: a single-institution discovery cohort (n=107) and a multi-institution validation cohort (n=497). We also serially examined clinical/pathological variables including time to disease recurrence. Created with BioRender.com

Specifically, when we compared *PCA3* expression in the validation cohort from TCGA, although average expression in all grades, stages, and groups was higher than in normal prostate, we found that among tumors there was significantly decreased *PCA3* expression in tumors with higher grades (Gleason score >7) and in higher stages (>pT2), as compared with lower grades, stages, or groups, respectively. These paradoxical findings are consistent with several early studies^17,18^ and in particular with a recent tissue-based study of *PCA3* expression in prostate cancer^19^. In that large cohort study, lower levels of tumor *PCA3* in both biopsy and radical prostatectomy specimens were associated with high-grade tumors, and in radical prostatectomy specimens decreased *PCA3* expression was associated with features of higher stages. Based on these results, it has been proposed that *PCA3* might actually represent a differentiation marker in human prostate cancer^19^. The finding of decreasing *PCA3* expression with increasing tumor grades and stages in both our study and others is broadly consistent with another previous study^20^, which found that the class of antisense intronic RNAs was markedly over-represented among the top transcripts associated with tumor differentiation in human prostate cancer. The finding of an inverse association between *PCA3* expression and increasing grades and stages may also relate to links between *PCA3* expression and androgen receptor (AR) signaling and the likelihood of *PCA3* having an important role in the early steps of prostate cancer carcinogenesis, with a reduced role when the disease is more advanced. Indeed, previous work by our own group and by others indicates that *PCA3* is upregulated by AR signaling^6-8^, and that *PCA3* is also involved in modulating AR signaling^7,21^. Interestingly, it also has been shown *in vitro* that *PCA3* silencing sensitizes prostate cancer cells to enzalutamide-induced decreased cell growth^22^. Alshalalfa et al. suggest that because low pretreatment serum testosterone levels are associated with diseases with higher grades and stages, and because of the relationship between AR signaling and *PCA3* expression, therefore lower *PCA3* expression may reflect the lower serum testosterone in these patients^19^. Although, we do not have any data on the pretreatment serum concentration of testosterone and other androgens, our tumor *PCA3* expression findings could perhaps be consistent with that interpretation.

Because prostate cancers, especially Gleason Score 7 (Groups 2 and 3) tumors, are quite frequent (about half of the total cases) and show divergent clinical behavior, there is great interest in developing prognostic biomarkers for risk stratification. Studies on the association of *PCA3* expression levels with outcome and prognosis show conflicting results^23^, and unlike this present study, most prior reports are based on urinary *PCA3* expression^24-26^. Our exploration of the validation cohort from TCGA, which comprised a wide spectrum of tumor grades and stages, revealed an association between lower levels of tumor *PCA3* expression and biochemical recurrence; however, this association was not found after taking grade and stage into account. This finding makes sense, as increasing grade and stage are both variables that are associated with lower *PCA3* expression. In their tissue-based cohort, Alshalalfa et al.^19^ also found an association between low *PCA3* levels and adverse outcomes, including biochemical recurrence, metastasis and prostate cancer-specific mortality; however, it is not clear whether such findings are independent of clinical and pathological variables (such as Gleason grade, stage, and group), as a multivariable analysis was not reported. For the discovery cohort of patients, we selected organ-confined, intermediate-risk tumors (Gleason groups 2 and 3, with tumor stages pT2 and pT3) where prognostic information might be expected to be most helpful clinically, to test for an association with outcome. We did not see any association between tumor *PCA3* expression and biochemical recurrence.

*PRUNE2*, a human homolog of the *Drosophila prune* gene, encodes for a protein with BCH, DHHA2, and PPX1 functional domains^7^. The BCH domain can inhibit the Rho family of proteins, small GTPases with roles in cell transformation, migration and metastasis, and cell cycle progression^5,27^. Evidence is accumulating that *PRUNE2* might act as a tumor suppressor gene. Loss-of-function mutations have been described in several tumor types, including germline and somatic mutations in parathyroid cancer^28^ and somatic mutations in solid papillary carcinoma^29^, while high expression of PRUNE2 protein correlates with favorable prognosis in neuroblastoma^30^. Others have shown evidence of inactivating *PRUNE2* mutations in Merkel cell carcinoma^31^ and that the restoration of downregulated PRUNE2 in oral cancer suppresses tumor cell migration^32^, further supporting the role of *PRUNE2* as a tumor suppressor. In prostate cancer, the evidence is limited and controversial: an early report found that *PRUNE2* expression was upregulated in prostate cancer and metastases in a small number of samples, and was androgen-inducible in prostate cancer cells^5^. However, a subsequent study on a larger number of samples found that *PRUNE2* expression either decreased or did not increase in aggressive prostate cancer, and that *PRUNE2* expression was not androgen-inducible^17^.

Altogether, the findings in the current study provide additional support for our previous findings^8^ that *PRUNE2* acts as a functional tumor suppressor gene in human prostate cancer. Here we described consistently lower expression of *PRUNE2* in prostate cancers of all grades and stages as compared to normal prostate. The findings in our present study are also consistent with the negative regulation of *PRUNE2* by *PCA3* in prostate cancer. We found no significant differences in *PRUNE2* expression across tumor stage, and only a small decrease in expression with increasing tumor grade, suggesting that loss of *PRUNE2* tumor suppressor activity is an early molecular event in prostate cancer. We are not aware of any prior reports of the prognostic significance of tumor *PRUNE2* expression in prostate cancer but, at least in this retrospective study of two independent prostate cancer patient cohorts, we did not find any association between *PRUNE2* expression and biochemical outcomes.

Strengths of this study include that broadly consistent findings were described in the two independent well-characterized clinically annotated primary prostate cancer cohorts used for analysis, and that the findings were robust across multiple assays in the discovery patient cohort and between the different methods of measurement of gene expression used in the two cohorts. The assessment of *PCA3* expression directly and specifically in tissue (as opposed to urine) is a novelty and a strength as our primary goal was the study of the *PRUNE2/PCA3* regulatory axis in human prostate cancer. We reasoned that the study of tissue expression is likely more informative of tumor biology than traditional urinalysis, not least of all because urinary expression, though very well characterized, could by subject to potential confounding issues such as RNA stability in urine or the contribution of differential urinary shedding. However, from the standpoint of assessment of prognostic information, a drawback of analyzing tissue *PCA3* expression is that the results are not directly comparable to the multiple previous studies that measured urinary *PCA3* scores and ultimately led to FDA and EMA approval for clinical applications in the US and EU. Moreover, while we did find consistent findings with a large tissue cohort study relating *PCA3* expression and biochemical recurrence^19^, the analysis presented here was limited in its ability to unequivocally determine the prognostic value of *PCA3* and *PRUNE2* expression as the overall proportion of patients with biochemical recurrences was relatively low. Finally, we were not able to fully address the relationship of reciprocal gene expression of *PCA3* and *PRUNE2* to the outcomes of metastases and prostate cancer-specific deaths, again due to the relative paucity of these events.

In conclusion, we found consistent upregulation of *PCA3* and downregulation of *PRUNE2* in prostate cancer as compared with normal prostate in two retrospective and independent patient cohorts (summarized in **Figure 4**), supporting that *PCA3* and *PRUNE3* function as an oncogene and a tumor suppressor gene, respectively, in human prostate cancer. The inverse correlation of *PCA3* and *PRUNE2* expression is consistent with our prior findings of a functional interplay between the two genes as part of a unique regulatory unit functioning at a single genetic locus in prostate cancer cells with *PCA3* negatively down-regulating *PRUNE2* expression^8^. The mechanistic dysregulation of *PCA3* and *PRUNE2* is observed across the spectrum of tumor grades and stages, suggesting that this is an early and stable molecular event in prostate cancer. On the other hand, we have not detected any regulatory effects of *PRUNE2/PCA3* in late genetic events such as prostate cancer progressing to biochemical recurrence, which includes the development of local tumor recurrence and/or the development of metastatic disease. The findings presented here represent additional evidence for the functional reciprocal co-regulation of *PCA3* and *PRUNE2* in the setting of early tumorigenesis but not in late events in human prostate cancer. Taken together along with the well-documented specificity of *PCA3* overexpression, our findings establish the *PCA3*/*PRUNE2* regulatory axis as an attractive early molecular target candidate for intervention in the therapy of human prostate cancer.

## Data Availability

For the discovery cohort, all data generated or analyzed are included in the manuscript and supporting files, except for patient-level ethnicity data. Patient-level ethnicity data is not included due to the potential for identifiability. However detailed summary ethnicity data is presented in the manuscript and in Table 1. Source codes are also available in the supplemental files. For the Validation Cohort, clinicopathological patient characteristics and gene level transcription data from The Cancer Genome Atlas (TCGA) were accessed from the UCSC Xena Resource.

https://xenabrowser.net/datapages/?dataset=TCGA.PRAD.sampleMap%2FHiSeqV2&host=https%3A%2F%2Ftcga.xenahubs.net&removeHub=https%3A%2F%2Fxena.treehouse.gi.ucsc.edu%3A443

## Author contributions

RCL, MB, EDN, DJM, RP, WA conceptualized the project; RCL, MB, TLS, AMT, JW, RD, JHL, AR, ASD, MAA, DNN, ITS, EDN, DJM collected and curated the data; RCL, MB, TLS, AMT, JW, RD, JHL, AR, ASD, MAA, DNN, ITS, EDN, DJM performed experiments and provided reagents or critical insights; RCL, MB, TLS, AMT, JW, RD, JHL, AR, ASD, MAA, DNN, ITS, EDN, IC, DJM, WKC, RP, WA analyzed the data; RCL, MB, EDN, DJM, RP, WA wrote the first manuscript draft to which all authors contributed edits; RCL, EDN, DJM, RP, WA obtained funding for the study. RCL, MB, RP, WA, supervised the study.

## Acknowledgements

We received support from the Human Tissue Repository and Tissue Analysis Shared Resource from the Department of Pathology of the University of New Mexico School of Medicine and a pilot award from the University of New Mexico Comprehensive Cancer Center. RP and WA were supported by awards from the Levy-Longenbaugh Donor-Advised Fund and the Prostate Cancer Foundation. EDN reports research fellowship support from the Brazilian National Council for Scientific and Technological Development (CNPq), Brazil, and the Associação Beneficente Alzira Denise Hertzog Silva (ABADHS), Brazil. This work has been funded in part by the NCI Cancer Center Support Grants (CCSG; P30) to the University of New Mexico Comprehensive Cancer Center (CA118100) and the Rutgers Cancer Institute of New Jersey (CA072720). The results shown here are in part based upon data generated by the TCGA Research Network: https://www.cancer.gov/tcga. These data have been presented in part at the 2017 American Society of Clinical Oncology Annual Meeting:

Richard C. Lauer, Marc Barry, Jin Wu, Ji-Hyun Lee, Dennis J. McCance, Ruofei Du, Arpit Rao, Andrey S. Dobroff, Emmanuel Dias-Neto, Isan Chen, Renata Pasqualini, and Wadih Arap. Evaluation of *PRUNE2* and *PCA3* expression as metastasis predictors in Gleason 7 prostate cancer. *J Clin Oncol* 2017; 35:15_suppl, e16583.

Richard C. Lauer, Marc Barry, Jin Wu, Ji-Hyun Lee, Dennis J. McCance, Ruofei Du, Arpit Rao, Andrey S. Dobroff, Emmanuel Dias-Neto, Isan Chen, Renata Pasqualini, and Wadih Arap. PRUNE2 and PCA3 expression in paired non-malignant and tumor specimens from radical prostatectomy patients with Gleason score 7 prostate cancer. *J Clin Oncol* 2017; 35:15_suppl, e16582.

Emmanuel Dias-Neto, Diana N. Nunes, Andrew M. Thomas, Tracey L. Smith, Arpit Rao, Richard C. Lauer, Isan Chen, Wadih Arap, Renata Pasqualini. *PCA3* upregulation in prostate cancer: Analysis in a cohort of 497 subjects from TCGA. *J Clin Oncol* 2017 35:15_suppl, e16578.

## Source Data

**Table 1 - Source Data File 1. Discovery Cohort**. Clinicopathologic features of the 107 patients in this study who had radical prostatectomy at UNM for localized prostatic adenocarcinoma

**Figure 1 - Source Data File 1. Analyses of discovery prostate cancer cohort**. Raw values of *PRUNE2* and *PCA3* expression in tumor prostatic samples.

**Figure 1 - Source Data File 2. Analyses of discovery prostate cancer cohort**. Raw values of *PRUNE2* and *PCA3* expression in nonneoplastic prostatic samples.

**Figure 1 - Source Data File 3. Analyses of discovery prostate cancer cohort**. Calculated values of *PRUNE2* and *PCA3* expression in tumor and nonneoplastic prostatic samples.

**Source Code File. R code and SAS code of descriptive statistics**.

## Supplemental Material

### Results

In the discovery cohort, to further assess the association between time to biochemical recurrence and the gene expression of *PRUNE2* and *PCA3* and the ratio of their expressions, and to take into account possible confounding by clinicopathological variables, we performed multivariate Cox proportional hazards regression modeling (raw data available in Supplemental File 1 tables and code). As there may be possible correlations between tumor Gleason Grade (GG) and tumor pathologic stage (PS), we created a composite categorical variable (“GG-PS”) representing the four possibilities in the discovery cohort: 3+4/pT2, 4+3/pT2, 3+4/pT3a, 4+3/pT3a. We then used two approaches to model outcome.

In the first approach, we fit four different Cox models. The explanatory variables in these models were as follows: model 1 (“GG_PS model”) – GG_PS only; model 2 (“*PRUNE2* model”) – *PRUNE2* expression, age, race, GG_PS, and interaction between GG_PS and *PRUNE2* expression; model 3 (“PCA3 model”) – *PCA3* expression, age, race, GG_PS, and interaction between GG_PS and *PCA3* expression; and model 4 (“Ratio model”) – *PRUNE2/PCA3* ratio, age, race, GG_PS, and interaction between GG_PS and the ratio. The goodness-of-fit of the models were compared using the Akaike Information Criterion (AIC), and the results are summarized in **Supplemental Table S1**. As a lower value of AIC indicates a better association fit, the model comparison indicates that the model with GG_PS only (model 1) represents the best fit for the data, and does not suggest that the expression of *PRUNE2, PCA3* or their ratio adds to the ability of pathology grade and stage to predict biochemical recurrence.

As a second multivariable approach to an assessment of the association of time to biochemical recurrence and gene expression, we created a multivariable model, including the following variables: expression of *PRUNE2*, expression of *PCA3*, age, race, the composite variable GG_PS, interaction between *PRUNE2* expression and GG_PS, and interaction between *PCA3* expression and GG_PS. In this case, a stepwise selection algorithm was used for model selection, and only the model with Gleason score and pathologic stage (GG_PS) was selected, as none of the other variables had a p-value less than the specified significance level of 0.25 (data not shown).

In the TCGA cohort, we used Cox proportional hazards modeling to assess the association of tumor PCA3 expression, adjusting for tumor grade, tumor stage, and age at diagnosis. The results are summarized in **Supplemental Table S2**. Briefly, on multivariable modeling, there was an association between tumor grade and stage with recurrence, we did not find that tumor *PCA3* expression was associated with biochemical recurrence [HR, 0.96; 95% CI 0.87-1.04, (p-value=0.36)].

### Methods

#### Statistical Analysis for Quantifying the expression of PCA3 and PRUNE2

There were combinations of assays and control genes used for quantifying the expression of *PCA3* and *PRUNE2* in this study. Explicitly, there were 9 duplex mixes for *PRUNE2*: PR1C1, PR1C2, PR1C3, PR2C1, PR2C2, PR2C3, PR3C1, PR3C2, PR3C3; and 6 duplex mixes for *PCA3*: PC1C1, PC1C2, PC1C3, PC2C1, PC2C2, PC3C3, where the first 3 letters denote an assay and last two letters denote a control gene being used in a particular run. For example, PC2C2 denotes the second assay for *PCA3* (Hs03462121_m1, detailed in Methods) and the second endogenous control gene (Hs02800695_m1, detailed in Methods) were used for that specific experiment. ⁏_⁏_is to denote the logarithmic number of PCR cycle when the fluorescent signal passes a threshold value. Let Δ⁏_⁏_ = ⁏_⁏ study gene_ – ⁏_⁏ control gene_ and we had –Δ⁏_⁏_ to quantify the gene expression (relative to a control gene), resulting in a positive value meaning an upregulated gene’s expression.

The experiment was completed 3 times for each gene duplex mix, e.g. we have three data points of PC2C2 measure for a tumor sample. The median of the three –Δ⁏_⁏_ values is summarized to estimate the gene expression of a particular gene duplex mix. We then looked at both mean and median of 9 estimates for *PRUNE2* and 6 estimates for *PCA3*, separately (data not shown). We did not see any significant difference utilizing mean or median in this or subsequent analyses.

### Supplemental Tables

**Supplementary Table S1.** Discovery cohort multivariable model

| Model | Akaike Information Criterion Score |
| --- | --- |
| PS_GG model | 157.2 |
| *PRUNE2* model | 162.9 |
| *PCA3* model | 166.2 |
| Ratio model | 162.2 |

**Supplementary Table S2.** Validation cohort multivariable cox model

|  | Hazard Ratio (HR) | 95% CI | p-value |
| --- | --- | --- | --- |
| *PCA3* | 0.963 | 0.888 – 1.044 | 0.36 |
| Gleason Grade | 1.558 | 1.147 – 2.117 | <0.01 |
| Pathologic Stage T3 | 3.596 | 1.360 – 9.512 | <0.01 |
| Pathologic Stage T4 | 1.860 | 0.206 – 16.82 | 0.58 |
| Age at Diagnosis | 1.000 | 0.960 – 1.043 | 0.99 |

### Supplemental Figures

**Supplementary Figure S1:**
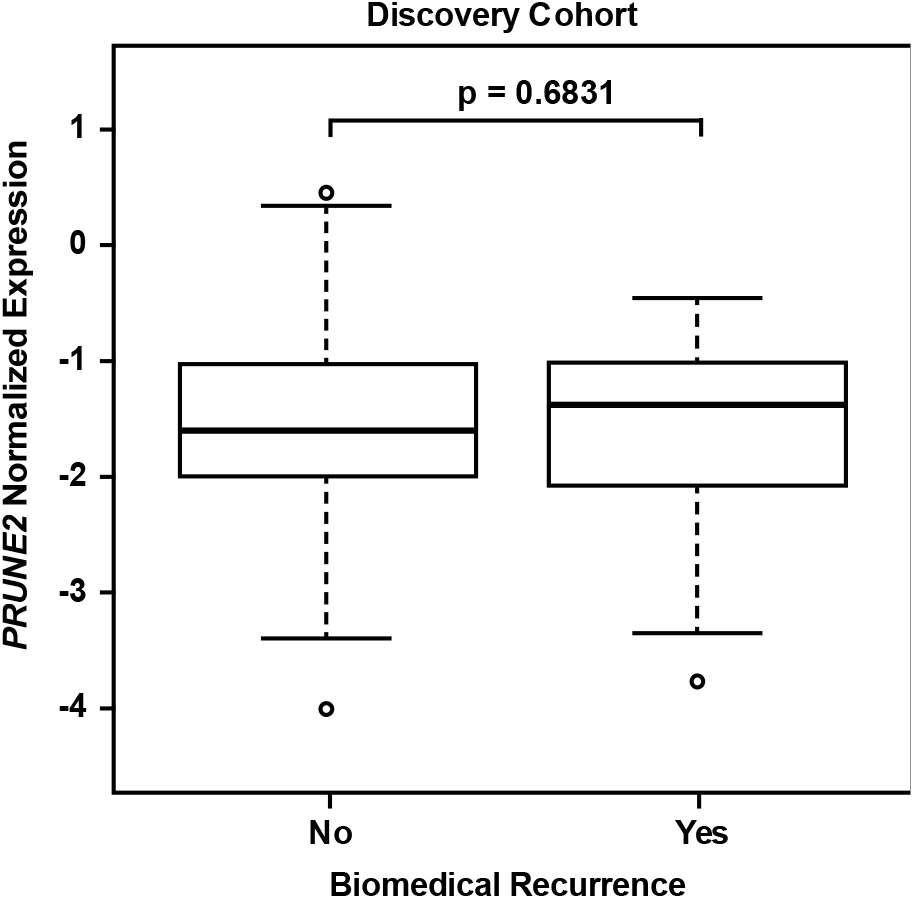
Discovery cohort - no significant difference in tumor *PRUNE2* expression by biochemical recurrence status.

**Supplementary Figure S2:**
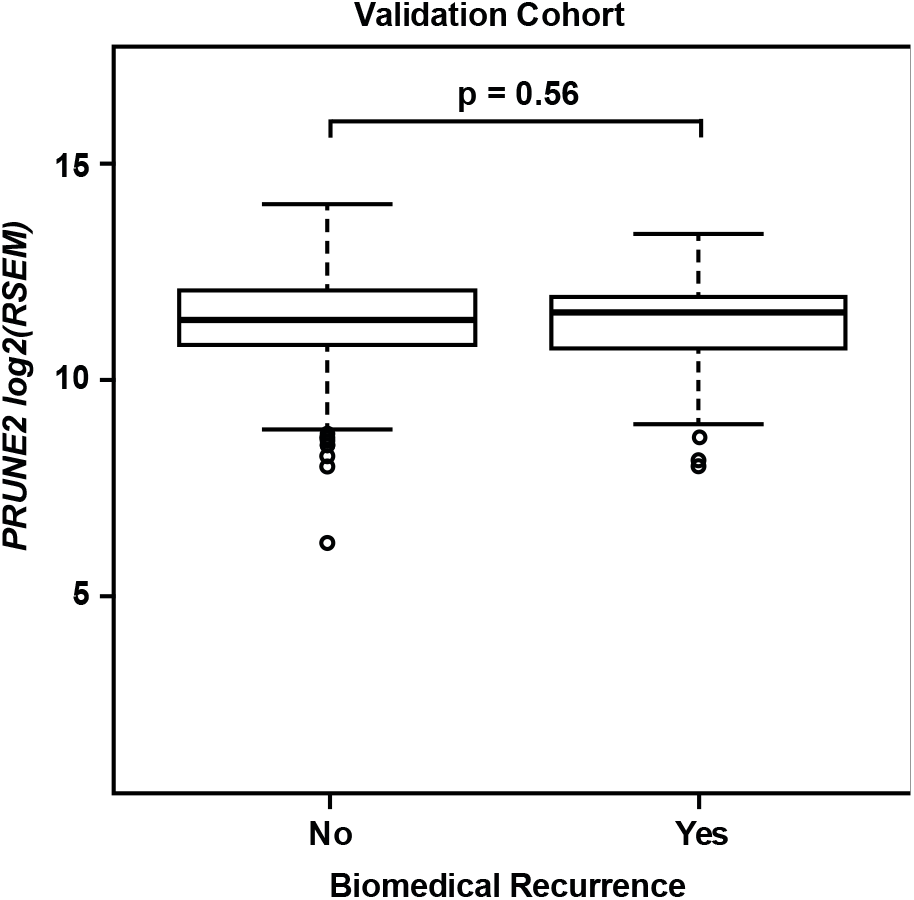
Validation Cohort – no significant difference in tumor *PRUNE2* expression by biochemical recurrence status.

**Table 1 - Supplemental Figure 1:**
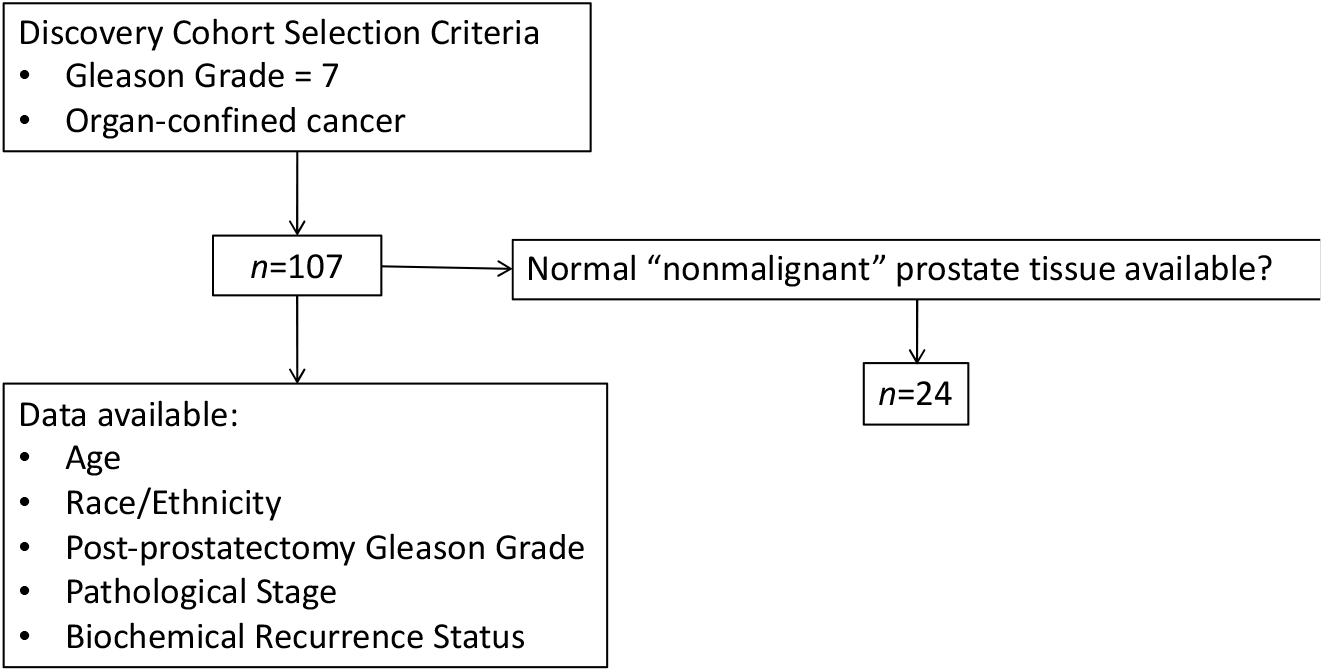
Flow diagram of Discovery Cohort selection criteria and clinicopathological characteristics available.

## References

1. Siegel RL, Miller KD, Fuchs HE, Jemal A. Cancer Statistics, 2021. CA Cancer J Clin. Jan 2021;71(1):7–33. doi:10.3322/caac.21654

2. Arun G, Diermeier SD, Spector DL. Therapeutic Targeting of Long Non-Coding RNAs in Cancer. Trends Mol Med. Mar 2018;24(3):257–277. doi:10.1016/j.molmed.2018.01.001

3. Bussemakers MJ, van Bokhoven A, Verhaegh GW, et al. DD3: a new prostate-specific gene, highly overexpressed in prostate cancer. Cancer Res. Dec 1 1999;59(23):5975–9.

4. de Kok JB, Verhaegh GW, Roelofs RW, et al. DD3(PCA3), a very sensitive and specific marker to detect prostate tumors. Cancer Res. May 1 2002;62(9):2695–8.

5. Clarke RA, Zhao Z, Guo AY, et al. New genomic structure for prostate cancer specific gene PCA3 within BMCC1: implications for prostate cancer detection and progression. PLoS One. 2009;4(3):e4995. doi:10.1371/journal.pone.0004995

6. Teixeira AA, Marchio S, Dias-Neto E, et al. Going viral? Linking the etiology of human prostate cancer to the PCA3 long noncoding RNA and oncogenic viruses. EMBO Mol Med. Oct 2017;9(10):1327–1330. doi:10.15252/emmm.201708072

7. Ferreira LB, Palumbo A, de Mello KD, et al. PCA3 noncoding RNA is involved in the control of prostate-cancer cell survival and modulates androgen receptor signaling. BMC Cancer. Nov 6 2012;12:507. doi:10.1186/1471-2407-12-507

8. Salameh A, Lee AK, Cardo-Vila M, et al. PRUNE2 is a human prostate cancer suppressor regulated by the intronic long noncoding RNA PCA3. Proc Natl Acad Sci U S A. Jul 7 2015;112(27):8403–8. doi:10.1073/pnas.1507882112

9. Gordetsky J, Epstein J. Grading of prostatic adenocarcinoma: current state and prognostic implications. Diagn Pathol. Mar 9 2016;11:25. doi:10.1186/s13000-016-0478-2

10. van Leenders G, van der Kwast TH, Grignon DJ, et al. The 2019 International Society of Urological Pathology (ISUP) Consensus Conference on Grading of Prostatic Carcinoma. Am J Surg Pathol. Aug 2020;44(8):e87–e99. doi:10.1097/PAS.0000000000001497

11. Livak KJ, Schmittgen TD. Analysis of relative gene expression data using real-time quantitative PCR and the 2(-Delta Delta C(T)) Method. Methods. Dec 2001;25(4):402–8. doi:10.1006/meth.2001.1262

12. Vandesompele J, De Preter K, Pattyn F, et al. Accurate normalization of real-time quantitative RT-PCR data by geometric averaging of multiple internal control genes. Genome Biol. Jun 18 2002;3(7):RESEARCH0034. doi:10.1186/gb-2002-3-7-research0034

13. Cancer Genome Atlas Research N. The Molecular Taxonomy of Primary Prostate Cancer. Cell. Nov 5 2015;163(4):1011–25. doi:10.1016/j.cell.2015.10.025

14. University of North Carolina TCGA Genome Characterization Center. TCGA prostate adenocarcinoma (PRAD) gene expression by RNAseq (polyA+ IlluminaHiSeq). 2017. https://xenabrowser.net/datapages/?dataset=TCGA.PRAD.sampleMap%2FHiSeqV2&host=https%3A%2F%2Ftcga.xenahubs.net&removeHub=https%3A%2F%2Fxena.treehouse.gi.ucsc.edu%3A443

15. Li B, Dewey CN. RSEM: accurate transcript quantification from RNA-Seq data with or without a reference genome. BMC Bioinformatics. Aug 4 2011;12:323. doi:10.1186/1471-2105-12-323

16. Goldman MJ, Craft B, Hastie M, et al. Visualizing and interpreting cancer genomics data via the Xena platform. Nat Biotechnol. 06 2020;38(6):675–678. doi:10.1038/s41587-020-0546-8

17. Salagierski M, Verhaegh GW, Jannink SA, Smit FP, Hessels D, Schalken JA. Differential expression of PCA3 and its overlapping PRUNE2 transcript in prostate cancer. Prostate. Jan 1 2010;70(1):70–8. doi:10.1002/pros.21040

18. Balcerczak E, Mirowski M, Sasor A, Wierzbicki R. Expression of p65, DD3 and c-erbB2 genes in prostate cancer. Neoplasma. 2003;50(2):97–101.

19. Alshalalfa M, Verhaegh GW, Gibb EA, et al. Low PCA3 expression is a marker of poor differentiation in localized prostate tumors: exploratory analysis from 12,076 patients. Oncotarget. Aug 1 2017;8(31):50804–50813. doi:10.18632/oncotarget.15133

20. Reis EM, Nakaya HI, Louro R, et al. Antisense intronic non-coding RNA levels correlate to the degree of tumor differentiation in prostate cancer. Oncogene. Aug 26 2004;23(39):6684–92. doi:10.1038/sj.onc.1207880

21. Lemos AE, Ferreira LB, Batoreu NM, de Freitas PP, Bonamino MH, Gimba ER. PCA3 long noncoding RNA modulates the expression of key cancer-related genes in LNCaP prostate cancer cells. Tumour Biol. Aug 2016;37(8):11339–48. doi:10.1007/s13277-016-5012-3

22. Ozgur E, Celik AI, Darendeliler E, Gezer U. PCA3 Silencing Sensitizes Prostate Cancer Cells to Enzalutamide-mediated Androgen Receptor Blockade. Anticancer Res. Jul 2017;37(7):3631–3637. doi:10.21873/anticanres.11733

23. Loeb S, Partin AW. Review of the literature: PCA3 for prostate cancer risk assessment and prognostication. Rev Urol. 2011;13(4):e191–5.

24. Loeb S, Bruinsma SM, Nicholson J, et al. Active surveillance for prostate cancer: a systematic review of clinicopathologic variables and biomarkers for risk stratification. Eur Urol. Apr 2015;67(4):619–26. doi:10.1016/j.eururo.2014.10.010

25. Lemos AEG, Matos ADR, Ferreira LB, Gimba ERP. The long non-coding RNA PCA3: an update of its functions and clinical applications as a biomarker in prostate cancer. Oncotarget. Nov 12 2019;10(61):6589–6603. doi:10.18632/oncotarget.27284

26. Fenstermaker M, Mendhiratta N, Bjurlin MA, et al. Risk Stratification by Urinary Prostate Cancer Gene 3 Testing Before Magnetic Resonance Imaging-Ultrasound Fusion-targeted Prostate Biopsy Among Men With No History of Biopsy. Urology. Jan 2017;99:174–179. doi:10.1016/j.urology.2016.08.022

27. Iwama E, Tsuchimoto D, Iyama T, et al. Cancer-related PRUNE2 protein is associated with nucleotides and is highly expressed in mature nerve tissues. J Mol Neurosci. Jun 2011;44(2):103–14. doi:10.1007/s12031-010-9490-2

28. Yu W, McPherson JR, Stevenson M, et al. Whole-exome sequencing studies of parathyroid carcinomas reveal novel PRUNE2 mutations, distinctive mutational spectra related to APOBEC-catalyzed DNA mutagenesis and mutational enrichment in kinases associated with cell migration and invasion. J Clin Endocrinol Metab. Feb 2015;100(2):E360–4. doi:10.1210/jc.2014-3238

29. Alsadoun N, MacGrogan G, Truntzer C, et al. Solid papillary carcinoma with reverse polarity of the breast harbors specific morphologic, immunohistochemical and molecular profile in comparison with other benign or malignant papillary lesions of the breast: a comparative study of 9 additional cases. Mod Pathol. Sep 2018;31(9):1367–1380. doi:10.1038/s41379-018-0047-1

30. Machida T, Fujita T, Ooo ML, et al. Increased expression of proapoptotic BMCC1, a novel gene with the BNIP2 and Cdc42GAP homology (BCH) domain, is associated with favorable prognosis in human neuroblastomas. Oncogene. Mar 23 2006;25(13):1931–42. doi:10.1038/sj.onc.1209225

31. Harms PW, Vats P, Verhaegen ME, et al. The Distinctive Mutational Spectra of Polyomavirus-Negative Merkel Cell Carcinoma. Cancer Res. Sep 15 2015;75(18):3720–3727. doi:10.1158/0008-5472.CAN-15-0702

32. Su SC, Yeh CM, Lin CW, et al. A novel melatonin-regulated lncRNA suppresses TPA-induced oral cancer cell motility through replenishing PRUNE2 expression. J Pineal Res. Oct 2021;71(3):e12760. doi:10.1111/jpi.12760

